# Intimate Partner Violence and Cancer Risk: A Systematic Review of Evidence and Gaps

**DOI:** 10.64898/2026.07.16.26358254

**Authors:** Dajana Glavas, Marie Anne Makoudjou, Giulia Melis, Lucia Bernardele, Nazareno Paolocci, Marco Scarpa, Jacopo Agrimi, Gaya Spolverato

## Abstract

**Background:** Despite its high prevalence and established impact on women’s health, the long-term biological effects of Intimate Partner Violence (IPV) remain poorly understood. In particular, its potential role in increasing cancer risk has received limited attention. This review examines whether IPV may be associated with elevated cancer risk in women.

**Methods:** We conducted a systematic review and meta-analysis in accordance with PRISMA and MOOSE guidelines to evaluate whether IPV may be associated with cancer risk. Eligible studies included adult women (≥18 years) with documented IPV exposure and cancer or precancerous outcomes. We searched PubMed, Web of Science, Scopus, and Google Scholar for articles published from 2000 to 2025. Study quality was assessed using the Newcastle–Ottawa Scale (NOS). A random-effects meta-analysis was performed on longitudinal studies reporting adjusted risk estimates.

**Results:** Thirteen studies were included in the qualitative synthesis, but only two met criteria for meta-analysis, both reporting on cervical cancer. The pooled odds ratio was 3.00 (95% CI: 2.05–4.38; I² = 0%). A separate pooled prevalence analysis of six retrospective studies showed that 32.2% of women with cancer reported a lifetime history of IPV. Study quality ranged from low to high.

**Conclusions:** This review underscores the limited and heterogeneous nature of the existing evidence on IPV as a potential cancer risk factor. While preliminary findings suggest a possible association, particularly with cervical cancer, the scarcity of high-quality longitudinal studies and the methodological variability in the studies reviewed prevent definitive conclusions regarding causal linkage. Further research, particularly prospective and mechanistic studies, is needed to clarify the relationship between IPV and oncogenesis across different cancer types and to identify underlying biological pathways.

## INTRODUCTION

Intimate Partner Violence (IPV) represents a pervasive and severe public health issue with significant global implications, particularly for women. Defined by the World Health Organization (WHO) as "any behavior within an intimate relationship that causes physical, psychological, or sexual harm",^1^ IPV encompasses physical, sexual, emotional, and psychological abuse by current or former partners.^2,3^ Although both men and women may experience IPV, women are disproportionately affected, facing serious physical and mental health consequences.^4,5^ Globally, one in three women has experienced physical and/or sexual IPV in their lifetime,^6^ and IPV is recognized as a leading cause of injury, chronic illness, and premature death among women worldwide.^4,7^

Beyond its immediate physical consequences, IPV has been associated with a broad spectrum of adverse health outcomes, including cardiovascular diseases, gastrointestinal disorders, respiratory illnesses, sexually transmitted infections, and profound mental health disorders such as depression, post-traumatic stress disorder (PTSD), and anxiety.^8–12^ IPV survivors also display increased rates of high-risk behaviors, including tobacco use, alcohol abuse, and illicit drug consumption, further compounding their health risks.^13,14^

Recent evidence suggests that IPV may also be implicated in cancer development, particularly gynecological cancers, raising the question of whether IPV constitutes a direct risk factor for oncogenesis.^15–18^ Epidemiological studies have documented significantly higher prevalence rates of cervical cancer among women with a history of IPV, in some cases up to ten times greater than those observed in the general population.^18,19^ IPV has been associated not only with cervical cancer but also with other tumor types, including breast cancer,^20^ although evidence remains more limited.

To date, the association between IPV and cancer has been largely attributed to indirect pathways, such as increased exposure to oncogenic infections (e.g., human papillomavirus, HPV), reduced adherence to cancer screening, and higher prevalence of behavioral risk factors like smoking and substance use.^21,22^ Socioeconomic status (SES) also plays a critical role, as it is closely intertwined with many of these factors, shaping access to healthcare, screening opportunities, and overall health behaviors, and may further exacerbate the impact of IPV on cancer development^20^. Nevertheless, emerging data suggest that IPV-related chronic stress may also influence cancer development through biological mechanisms, including immune dysregulation, chronic inflammation, and hormonal imbalances.^23,24,25^ Preclinical studies have demonstrated alterations in estrogen receptor beta (ERβ) expression and immune system function in models of chronic stress and IPV, pointing to possible molecular links between IPV and oncogenesis.^26^ Despite these insights, no comprehensive analysis has yet addressed whether IPV can be considered a direct and independent risk factor for cancer beyond its indirect effects.

To address this gap, we conducted a comprehensive review and meta-analysis aimed at systematically evaluating the association between IPV and cancer. By integrating both longitudinal studies investigating cancer incidence among IPV survivors and retrospective studies assessing the prevalence of IPV among cancer patients, we sought to explore whether IPV may represent a direct risk factor for cancer development. In addition to synthesizing available epidemiological evidence, we examined potential biological mechanisms underlying this association, including chronic inflammation, immune dysregulation, and hormonal alterations. This approach not only allows for a critical evaluation of current data but also highlights important implications for research, clinical practice, and public health strategies targeting the long-term health consequences of IPV.

## METHODS

### Reporting Guidelines

This systematic review and meta-analysis was conducted in accordance with the PRISMA (Preferred Reporting Items for Systematic Reviews and Meta-Analyses)^27^ and MOOSE (Meta-analysis of Observational Studies in Epidemiology)^28^ guidelines.

### Eligibility Criteria

Eligibility criteria were defined using the PECO framework. The population of interest consisted of adult women (aged 18 years or older) who had experienced Intimate Partner Violence (IPV), including physical, sexual, or psychological abuse. The exposure was defined as a history of IPV in adulthood. The comparison group included women with no history of IPV exposure. The primary outcome was the diagnosis of any form of cancer, or the presence of cancer-related abnormalities or precancerous lesions defined as cellular abnormalities that can progress to invasive cancer if left untreated^29^, such as high-risk HPV infection, abnormal Pap tests, or cervical intraepithelial neoplasia.

We included peer-reviewed primary studies published in English between 2000 and 2025, encompassing cross-sectional, case-control, cohort, prospective, and retrospective designs. We excluded editorials, reviews, commentaries, case reports, studies on pediatric or animal populations, and studies with small sample sizes (fewer than 10 participants).

### Search Strategy

A comprehensive literature search was conducted in four international databases: PubMed, Web of Science, Scopus, and Google Scholar.

Given Google Scholar’s relevance-based ranking algorithm^30^ and the high volume of non-peer-reviewed records in unrefined searches, we limited the screening to the first 500 records (50 pages). This threshold ensures a rigorous yet focused identification of relevant literature while systematically minimizing ’noise’^31^. This approach aligns with established evidence synthesis practices, which recognize that searching beyond the initial several hundred results typically yields diminishing returns^32^.

The search covered the period from January 2000 to March 2025, and the last search was completed on March 20, 2025. Search terms included both free-text and Medical Subject Headings (MeSH), combining terms for IPV (“intimate partner violence”, “IPV”, “domestic violence”, “spouse abuse”) and cancer (“cancer”, “neoplasm”, “malignancy”, “tumor”).

The search string used was:

*("Intimate Partner Violence"[Mesh] OR "Spouse Abuse"[Mesh] OR "Intimate Partner Violence"[All Fields] OR "Domestic Violence"[All Fields] OR "Spouse Abuse"[All Fields] OR "IPV"[All Fields]) AND ("Neoplasms"[Mesh] OR cancer*[All Fields] OR neoplasm*[All Fields] OR carcinoma*[All Fields] OR malignan*[All Fields] OR tumor*[All Fields] OR tumour*[All Fields])*

A total of 1392 articles identified in an initial analysis were thereafter screened on full text analysis. All articles reporting data on IPV and cancer diagnosis, epidemiology or outcome, and on possible mechanisms explaining this connection, were considered for review. Three reviewers (DG, MM and LB) independently screened titles and abstracts, and full texts were retrieved for all potentially eligible studies. Disagreements were resolved through discussion with a fourth reviewer (JA). Relevant data about Authors, year of publication, Journal, study design, sample size, cancer types and sizes and measures as incidence, prevalence, OR, aOR, relative risk (RR), or adjusted relative risk (aRR) were extracted for each paper.

### Study Selection

Following the initial screening, we performed an additional selection step to refine the scope of the meta-analysis. Our objective was not to infer directly causal linkage, but to identify studies that explicitly examined IPV as a potential contributing factor in cancer development. We therefore included only studies designed to assess whether prior exposure to IPV was associated with increased vulnerability to cancer through biological, behavioral, or psychosocial pathways. Eligible studies were either longitudinal in design and investigated the prospective association between IPV and cancer incidence, or retrospective/cross-sectional studies among women with cancer that evaluated IPV history as a possible etiological factor rather than a coincidental or post-diagnosis variable. For these latter studies, a temporal relationship where IPV exposure occurred before the cancer diagnosis was required for inclusion. This criterion aimed to exclude studies in which a temporal ordering could not be established, thereby ensuring at least the necessary, though not sufficient, conditions for considering a potential causal linkage between IPV exposure and cancer. This approach was intended to isolate the subset of studies most relevant to the hypothesis that IPV may function as an independent risk factor for cancer. A total of 13 studies met these criteria and were included in the qualitative synthesis (Fig.1).

**Figure 1:**
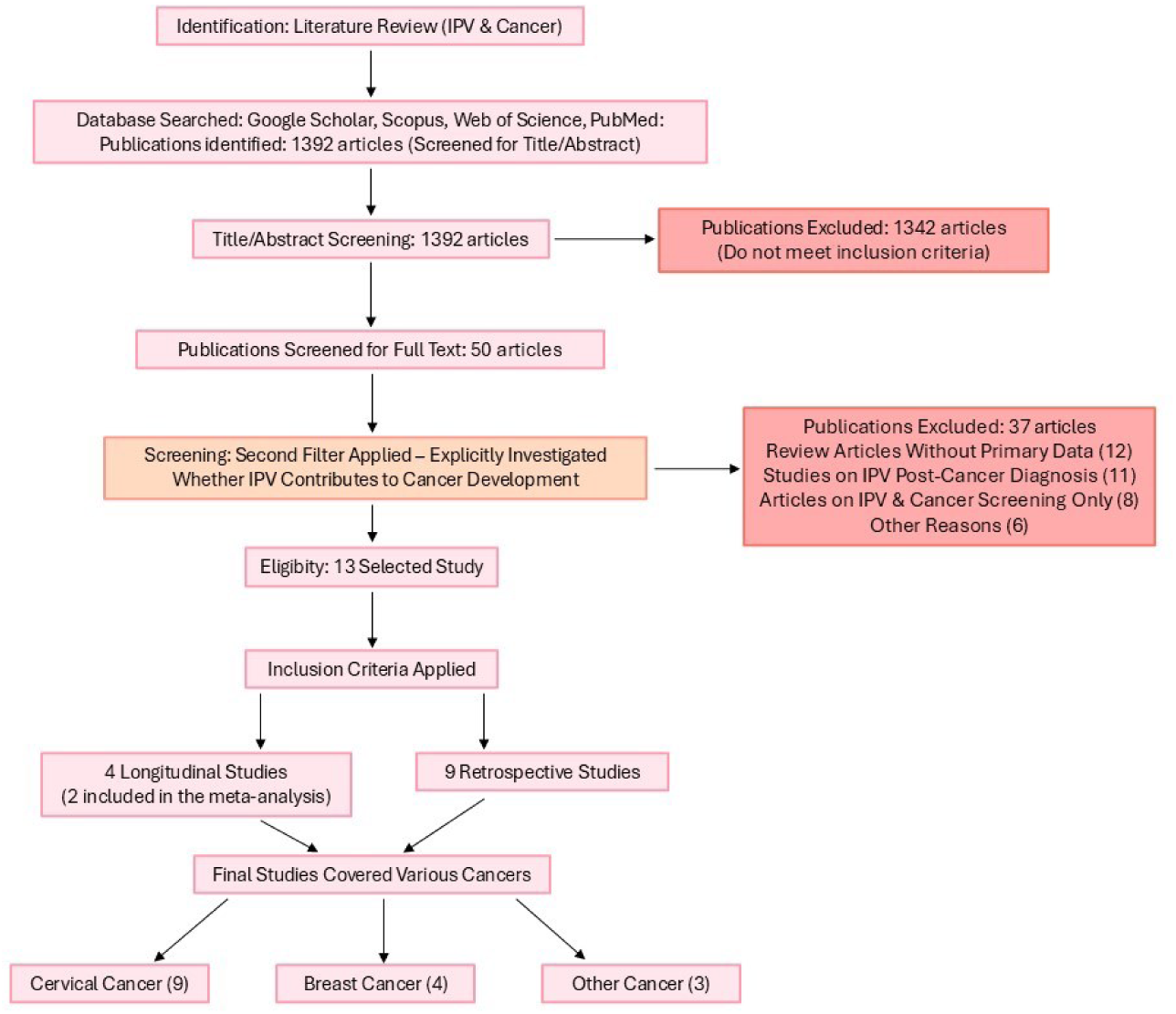
Flowchart of study selection process. Flow diagram illustrating the identification, screening, eligibility assessment, and inclusion of studies according to PRISMA guidelines. A total of 13 studies met the inclusion criteria and were included in the qualitative synthesis.

### Risk of Bias Assessment

We assessed the methodological quality of all studies using the Newcastle-Ottawa Scale (NOS). This tool evaluates three domains: selection of participants (maximum 4 points), comparability of study groups (maximum 2 points), and ascertainment of exposure or outcome (maximum 3 points). Each study was independently scored by two reviewers, with disagreements resolved by consensus. Scores were interpreted as follows: high quality (7–9), moderate quality (4–6), and low quality (0–3). Among the thirteen studies included in our analysis, five were rated as high quality (Coker et al., 2000^15^, Coker et al., 2017 A^33^, Jetelina et al., 2020^20^, Coker et al., 2017 B^34^, Coker et al., 2012^35^) and the others as moderate. Full itemized scoring is reported in Supplementary Table 1.

### Pooled Prevalence Analysis

A pooled prevalence analysis was conducted to estimate the prevalence of intimate partner violence (IPV) among women diagnosed with cancer, based on retrospective studies. Only studies explicitly reporting raw prevalence data of IPV among cancer patients were included in this analysis. Only six retrospective studies met the inclusion criteria for the prevalence analysis, all of which reported IPV prevalence among women already diagnosed with cancer. Studies were excluded if they lacked clear prevalence data, relied solely on high-risk HPV as an outcome, or failed to establish the temporal direction of IPV relative to cancer diagnosis. In contrast, studies reporting formal risk estimates such as odds ratios (OR) or adjusted odds ratios (aOR) were not included here, as those were analyzed separately in the meta-analysis section. Relevant data were extracted and organized into structured tables, including study design, cancer type, sample size, and epidemiological measures such as prevalence, incidence, OR, aOR, relative risk, or adjusted relative risk. Prevalence and incidence values were calculated from the available study data when explicit figures were not provided. All statistical analyses were performed using R software.

### Meta-analysis

Among the 13 included studies, only two provided formal risk estimates with 95% confidence intervals. These were included in the meta-analysis. Studies that provided only descriptive data, prevalence estimates, or qualitative outcomes were excluded from the quantitative synthesis but retained in the narrative analysis. A random-effects model (DerSimonian and Laird method) was used to account for heterogeneity across studies.

One of the two study (Coker et al., 2000)^15^ reported an adjusted relative risk (aRR). Given that the outcome under investigation—cervical cancer—is relatively rare in the study population, the aRR was considered sufficiently comparable to ORs to be included in the meta-analysis. This approach has been adopted in previous meta-analyses on violence and health outcomes when the outcome is uncommon and the difference between effect measures is expected to be minimal^36,37^. A sensitivity analysis was performed to assess the impact of including this study on the overall pooled estimate.

Each effect size was log-transformed to stabilize variance, and standard errors were calculated from the reported confidence intervals. Heterogeneity was assessed using both the Q statistic and the I² index, with thresholds of 25%, 50%, and 75% interpreted as low, moderate, and high heterogeneity, respectively.

### Additional Analyses Considered

Several advanced meta-analytic techniques were considered but not conducted due to the limited number of eligible studies (n=2) providing formal risk estimates. Specifically, we considered sensitivity analyses (e.g., leave-one-out procedures to assess robustness), subgroup analyses (e.g., stratifying by cancer type or study design), meta-regressions to explore potential moderators (e.g., study year, population characteristics), and publication bias assessments (e.g., funnel plot asymmetry tests). However, these analyses typically require a minimum number of studies (at least 10) to yield meaningful and interpretable results. Therefore, these analyses were not performed to avoid unreliable or misleading conclusions.

## RESULTS

### Study Selection and Characteristics

A total of thirteen studies met the predefined inclusion criteria and were included in this review. All selected studies were analyzed to evaluate the association between intimate partner violence (IPV) and cancer, considering both the prevalence of IPV among cancer patients and the prevalence of cancer among IPV-exposed populations, as well as broader qualitative aspects of this relationship. In addition to a comprehensive qualitative synthesis, we conducted pooled prevalence analyses based on data extracted from ten of the included studies to estimate both the occurrence of cancer in IPV survivors and the occurrence of IPV among women with cancer. Two studies by Coker et al. (2012^35^ and 2017 B^34^) were excluded from the pooled analyses due to overlapping populations. Only the most recent and comprehensive dataset by Coker et al. (2017 A^33^) was retained. In addition, two studies provided formal risk estimates (such as odds ratios or adjusted risk ratios) and were included in a dedicated meta-analysis of effect size. The selected studies primarily investigated cervical and breast cancer, with a predominance of cross-sectional and retrospective designs. However, there was considerable heterogeneity in study quality, particularly in terms of adjustment for major confounding factors, such as HPV infection, smoking, socioeconomic status, and sexual behavior. The characteristics of the included studies, stratified into longitudinal and retrospective designs, are summarized in Tables 1 and 2, respectively. For each study, we extracted detailed information including first author, year of publication, journal, study design, type of cancer investigated, sample size, and epidemiological data such as incidence, prevalence, and measures of association when available.

**Table 1.** Characteristics of included studies: Longitudinal studies. The table details study authorship, year, journal, design, total sample size, cancer types considered, and cancer incidence. When reported, both total and cancer-specific incidence rates are provided.

| First Author | Year | Journal | Study Design | Sample Size (n) | Cancer Type(s) and Size | Reported Cancer Incidence (%) |
| --- | --- | --- | --- | --- | --- | --- |
| Cesario | 2014 | Clin J Oncol Nurs. | *Interview-based cross-sectional* | 300 | Cervical (6); Thyroid cancer (1); Melanoma (1) | Cervical 2%; Thyroid 0·33%; Melanoma 0·33% |
| Coker | 2000 | J Womens Helath Gend Based Med | *Cross-sectional* | 1152 | Cervical | 1·2%; |
| Coker | 2009 | J Womens Health (Larchmt) | *Cross-sectional, registry* | 4732 | Cervical | 2·10%; |
| Madding | 2024 | BMC Womens Health | Observational | 16 | Cervical High-risk HPV | 18·75% |

**Table 2.**
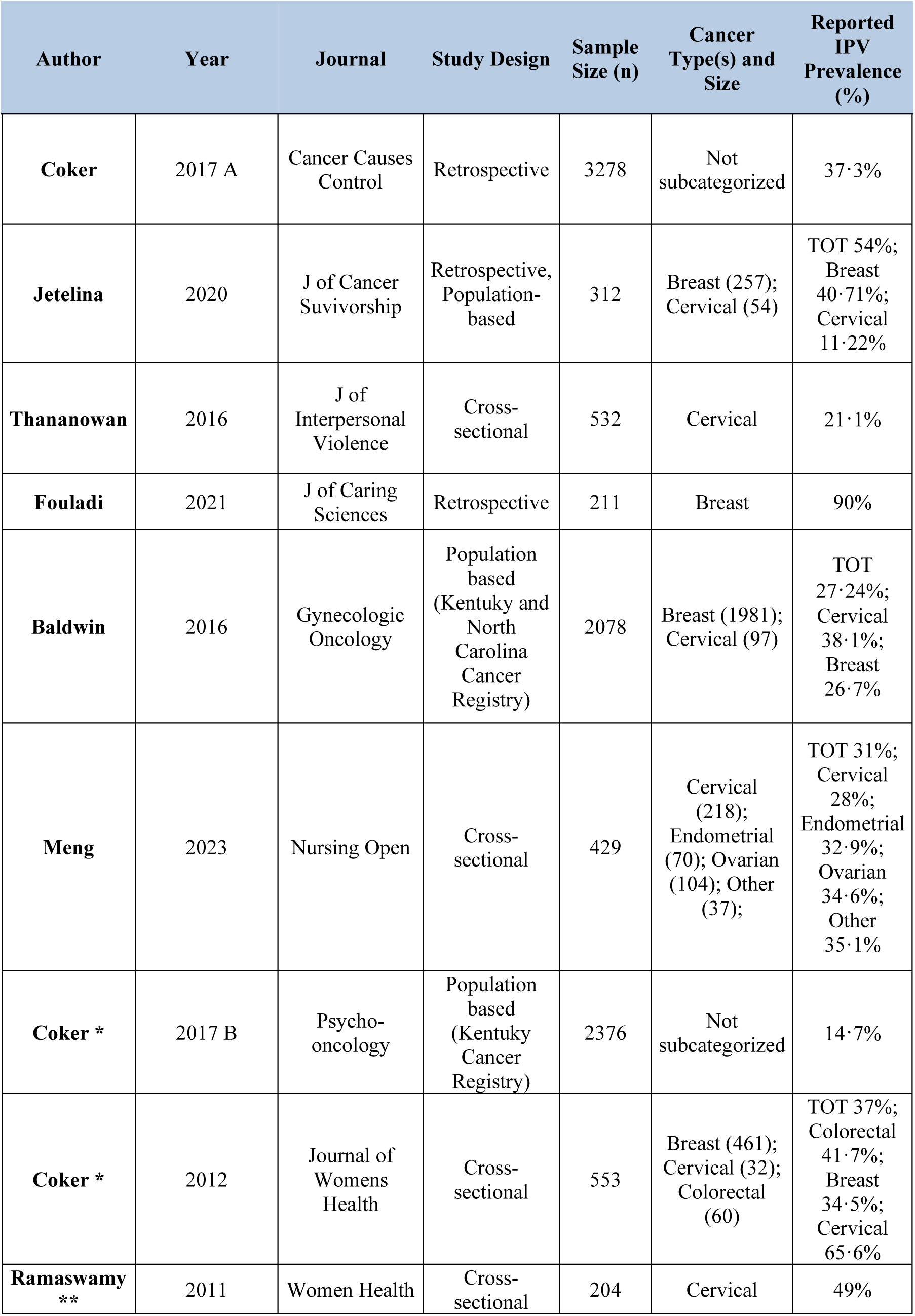
Characteristics of included studies: Retrospective studies. * studies excluded from pooled analysis due to overlapping population ** study excluded from pooled analysis due to unclear temporal directionality The table details study authorship, year, journal, design, total sample size, cancer types considered, and IPV prevalence. When reported, both total and cancer-specific IPV prevalence rates are provided

| Author | Year | Journal | Study Design | Sample Size (n) | Cancer Type(s) and Size | Reported IPV Prevalence (%) |
| --- | --- | --- | --- | --- | --- | --- |
| **Coker** | 2017 A | Cancer Causes Control | Retrospective | 3278 | Not subcategorized | 37·3% |
| **Jetelina** | 2020 | J of Cancer Survivorship | Retrospective, Population-based | 312 | Breast (257); Cervical (54) | TOT 54%; Breast 40·71%; Cervical 11·22% |
| **Thananowan** | 2016 | J of Interpersonal Violence | Cross-sectional | 532 | Cervical | 21·1% |
| **Fouladi** | 2021 | J of Caring Sciences | Retrospective | 211 | Breast | 90% |
| **Baldwin** | 2016 | Gynecologic Oncology | Population based (Kentucky and North Carolina Cancer Registry) | 2078 | Breast (1981); Cervical (97) | TOT 27·24%; Cervical 38·1%; Breast 26·7% |
| **Meng** | 2023 | Nursing Open | Cross-sectional | 429 | Cervical (218); Endometrial (70); Ovarian (104); Other (37); | TOT 31%; Cervical 28%; Endometrial 32·9%; Ovarian 34·6%; Other 35·1% |
| **Coker \*** | 2017 B | Psycho-oncology | Population based (Kentucky Cancer Registry) | 2376 | Not subcategorized | 14·7% |
| **Coker \*** | 2012 | Journal of Womens Health | Cross-sectional | 553 | Breast (461); Cervical (32); Colorectal (60) | TOT 37%; Colorectal 41·7%; Breast 34·5%; Cervical 65·6% |
| **Ramaswamy \*\*** | 2011 | Women Health | Cross-sectional | 204 | Cervical | 49% |

### Analysis and description of included studies

The longitudinal cohort included studies that examined patients with a history of IPV who later developed cancer, thereby capturing the forward progression of events from IPV exposure to cancer diagnosis. The research involved a total of 6,200 participants across four studies, investigating the association between IPV and an increased risk of cancer. Among these, three studies focused primarily on cervical cancer (Coker et al., 2000;^15^ Coker et al., 2009;^16^ Madding et al., 2024^38^), while additional tumor types, including melanoma and thyroid cancer, were examined in a smaller subset of participants (Cesario et al., 2014^19^).

In the prospective study by Cesario et al.^19^, 300 women with a history of IPV were assessed for cancer diagnoses and functional outcomes. Cervical cancer emerged as the most frequent diagnosis, with an incidence of 2%, with a tenfold higher prevalence compared to the general population. The study also identified isolated cases of thyroid cancer and melanoma (each with an incidence of 0**·**33%), suggesting that the oncogenic impact of IPV might extend beyond gynecological malignancies.

Coker et al.(2000)^15^ analyzed data from 1,152 women screened for IPV in two family practice clinics from 1997 till 1999, finding that 1**·**2% of participants reported a cervical cancer diagnosis, and 20**·**3% had undergone treatment for cervical neoplasia. Importantly, IPV was associated with a significantly increased risk of invasive cervical cancer (adjusted relative risk [aRR] 4**·**28; 95% CI: 1**·**94–18**·**39) and preinvasive cervical neoplasia (aRR 1**·**47; 95% CI: 1**·**16–1**·**82), with the strongest associations observed in women exposed to physical or sexual IPV, as opposed to psychological violence alone.

In a subsequent large-scale study on a different cohort of patients, Coker et al^16^ (2009) examined 4,732 women participating in the Kentucky Women’s Health Registry from 2006 till 2007, of whom 35**·**9% reported a history of IPV. Cervical cancer prevalence was 2**·**1% in the overall cohort but significantly higher (3**·**5%) among women who had experienced IPV or forced sex by non-partners, further supporting a link between violence exposure and cervical oncogenesis.

Finally, Madding et al.^38^ explored the impact of IPV on adherence to cervical cancer screening programs. In this observational study, women with a history of IPV demonstrated markedly lower screening adherence and higher rates of cervical dysplasia and cancer compared to general populations, highlighting the potential role of IPV in delaying diagnosis and increasing cancer risk through reduced access to preventive care.

The retrospective cohort included studies that identified patients with a cancer diagnosis and then retrospectively investigated their history for IPV exposure that occurred prior to the diagnosis. This cohort consisted of nine studies that investigated cancer patients with a history of IPV. These studies explored the association between violence exposure and various cancer types, including breast, cervical, endometrial, ovarian, vulvar, and colorectal cancers. These studies focused not only on the prevalence of IPV among women with cancer but also on how IPV may influence cancer subtypes, clinical outcomes, and access to care.

Four studies specifically addressed the relationship between IPV and breast cancer. In a population-based analysis, Jetelina et al.^20^ found that breast cancer patients with a history of IPV were twice as likely to develop estrogen receptor-negative (ER-) and/or progesterone receptor-negative (PR-) tumors, which are typically associated with worse prognoses. Moreover, IPV-exposed women were less likely to receive surgery and hormone therapy. In this study, IPV prevalence reached 40·71% among 257 breast cancer patients. Similarly, Fouladi et al.^39^ reported an exceptionally high IPV prevalence of 90% in a sample of 211 Iranian breast cancer patients, suggesting that IPV may be a pervasive but underrecognized factor in this population. Baldwin et al.^40^, examining 1981 women with breast cancer identified a 26·7% IPV prevalence. Although cervical and vulvar cancers were linked to an increased probability of IPV compared with breast cancer, IPV showed a crucial impact on cancer-related quality of life. Similarly, Coker et al.^35^ (2012) in a cross-sectional study on 461 breast cancer patients, underlined a 34·5% of IPV prevalence.

Regarding gynecological cancers, several studies analyzed the role of IPV in influencing cervical cancer risk and screening behaviors. Ramaswamy et al.^41^, in a cohort of 204 incarcerated women, found that those with a recent history of IPV were significantly more likely to report abnormal Pap smears (OR 2·41; 95% CI: 1·09–5·31), have been diagnosed with cervical cancer (OR 2·89; 95% CI: 0·90-9·27) and to have received treatment for cervical cancer (OR 5·44; 95% CI: 1·05–28·06), highlighting the severe health implications of IPV in vulnerable populations. Thananowan et al.^42^ similarly observed among a cohort of 532 Thai women that 21·1% reported IPV and 22.2% had cervical cancer, underscoring a potentially strong correlation between violence exposure and oncogenesis. They noted a significant association between IPV, psychological distress, and reduced social support, emphasizing the intersection between IPV and psychosocial determinants of health. Baldwin et al.^40^ provided further insights into the relationship between IPV and cancer subtypes, reporting a significantly higher prevalence of IPV among women with cervical or vulvar cancer (57.3%) compared to those with breast cancer (26·7%; p =·0003). Furthermore, women with cervical cancer exhibited higher rates of both past IPV (38·1% vs. 26·7%) and current IPV or partner interfering behavior (PIB) (18·6% vs. 10·2%), suggesting that IPV may play a specific role in the etiology or progression of certain gynecological cancers. Interestingly, no significant differences in IPV prevalence were observed among women with endometrial or ovarian cancer compared to breast cancer patients, and lifetime IPV or PIB were not associated with more advanced cancer stages. Finally, Meng et al.^43^ reported that 31% of 429 Chinese women with gynecological cancers had experienced IPV, although no association was found between IPV exposure and cancer stage or treatment pathways, pointing to a complex interplay of factors that warrants further investigation.

### Meta-Analysis Findings

Only two studies met the criteria for inclusion in the meta-analysis, each providing formal risk estimates (adjusted relative risk, aRR, or adjusted odds ratio, aOR) with 95% confidence intervals (95% CI) regarding the association between Intimate Partner Violence and cervical cancer (Table 3). Both studies focused specifically on cervical cancer and used large, well-defined samples. Coker et al.^15^ (2000) reported an adjusted relative risk of 4.28 (95% CI: 1.94–18.39), based on a retrospective cohort design with temporal verification of IPV exposure preceding cancer diagnosis. Coker et al.^16^ (2009) provided an adjusted odds ratio of 2.7 (95% CI: 1.8–4.0), derived from a large population-based registry.

**Table 3.** Characteristics of the studies included in the meta-analysis. The table details study authorship, year, journal, design, total sample size, cancer types considered, and reported cancer epidemiology detailed in Odds Radio (OR) or adjusted Relative Risk (aRR) with 95% Confidence Interval (CI).

| Author | Year | Journal | Study Design | Cancer Type (n) | Sample Size (n) | Reported Cancer Incidence/Prevalence |
| --- | --- | --- | --- | --- | --- | --- |
| Coker | 2000 | J Womens Health Gend Based Med | *Cross-sectional* | Cervical | 1152 | 1.2%; aRR 4.28 (95% CI: 1.94–18.39) |
| **Coker** | 2009 | J Womens Health (Larchmt) | ***Cross-sectional Registry-based*** | Cervical | 4732 | 2.1%; aOR 2.7 (95% CI: 1.8–4.0) |

Although the included studies employed different measures of association (aRR vs aOR), these estimates were considered comparable given the low incidence of cervical cancer and the use of adjusted values. Both studies explicitly confirmed the temporal sequence of IPV exposure preceding cancer, allowing inclusion in the meta-analysis.

The pooled analysis, conducted using a random-effects model, indicated that women with a history of IPV had an increased risk of cervical cancer, with a pooled OR of 3.00 (95% CI: 2.05–4.38) (Figure 2). Heterogeneity across studies was null (I² = 0%), indicating a consistent effect size.

**Figure 2.**
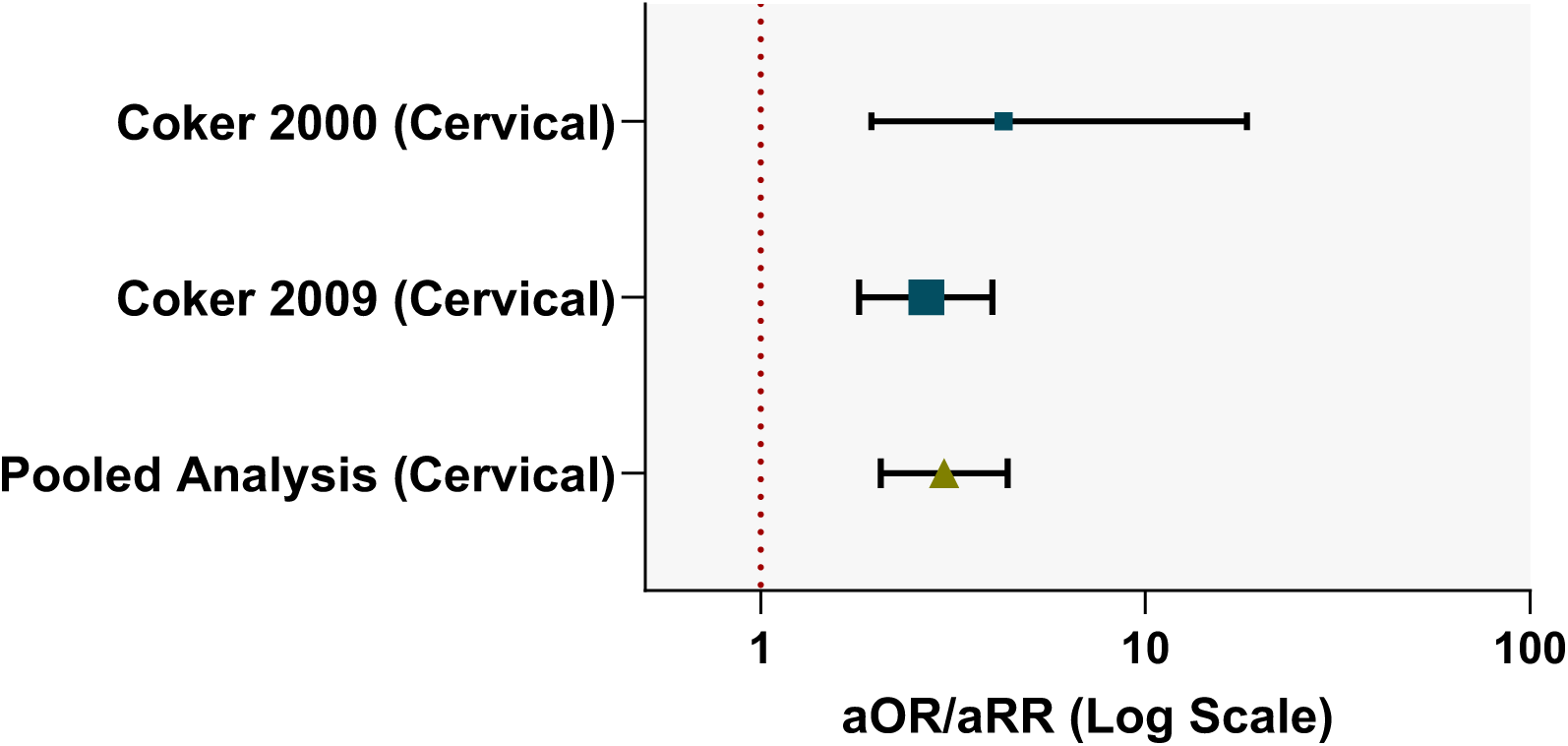
Forest plot of the meta-analysis assessing the association between Intimate Partner Violence (IPV) and cervical cancer. The meta-analysis included two studies (Coker et al., 2000; Coker et al., 2009) reporting adjusted risk estimates. The pooled odds ratio (OR) was 3.00 (95% CI: 2.05–4.38). Heterogeneity was low (I² = 0%).

A critical analysis of confounder adjustments revealed that neither of the studies included in the meta-analysis accounted for HPV infection, despite its established role in cervical carcinogenesis. Although HPV was acknowledged as a potential mediator in both studies, no biological or behavioral HPV-related variables were measured or adjusted for. Smoking was explicitly adjusted for in both studies, and SES was accounted for using proxies such as education and insurance coverage. Sexual behavior variables, including age at first intercourse and number of sexual partners, were only addressed by Coker et al.^15^ (2000). Although sexually transmitted infections (STIs) were discussed and partially examined through stratified analysis, they were not included as covariates in the final models. Other common covariates included age, marital status, and race/ethnicity. A summary of confounder adjustments is provided in Table 4.

**Table 4.** Confounding Factors Considered in Included Studies. The table details for each study the type of adjustment for confounding factors: Human Papilloma Virus (HPV), smoking, socioeconomic status (SES), sexual behavior, Sexually Transmitted Infections (STI) and other adjustments such as age, race, marital status, drug use.

| Study (Author, Year) | HPV Adjusted? | Smoking Adjusted? | SES Adjusted? | Sexual Behavior Adjusted? | STI Adjusted? | Other Adjustments | Notes |
| --- | --- | --- | --- | --- | --- | --- | --- |
| **Coker et al. (2000)** | No | Yes | Yes | Yes | No (considered but not adjusted) | Age, Medicaid, race, marital status | HPV discussed but not measured |
| **Coker et al. (2009)** | No | Yes | Yes | Partial | No | Age, drugs, marital status, race | HPV not measured, STI excluded |

### Pooled Prevalence Analysis findings

To provide a clearer picture of the bidirectional relationship between IPV and cancer, we conducted a pooled prevalence analysis focusing on retrospective studies. Six studies involving a total of 6,840 women with a cancer diagnosis were included, of whom 2,204 reported a lifetime history of IPV, yielding a pooled prevalence of 32.2% (Fig. 3, Tab. 5). Subgroup analyses showed a prevalence of 28.3% among women with breast cancer (n=2,601), 25.5% in those with cervical cancer (n=961), and 37.3% in studies where cancer type was not specified (n=3,278). Three studies were excluded from the pooled analysis: Coker^34^ (2017 B) and Coker^35^ (2012) due to overlapping cohorts, and Ramaswamy 2011 due to unclear temporal directionality. Given the limited number of longitudinal studies (n=4) and the substantial overlap with those included in the meta-analysis (notably, Coker et al., 2000^15^ and 2009^16^), we did not conduct a pooled prevalence analysis for this group, to avoid redundancy and potential inflation of shared findings.

**Fig. 3.**
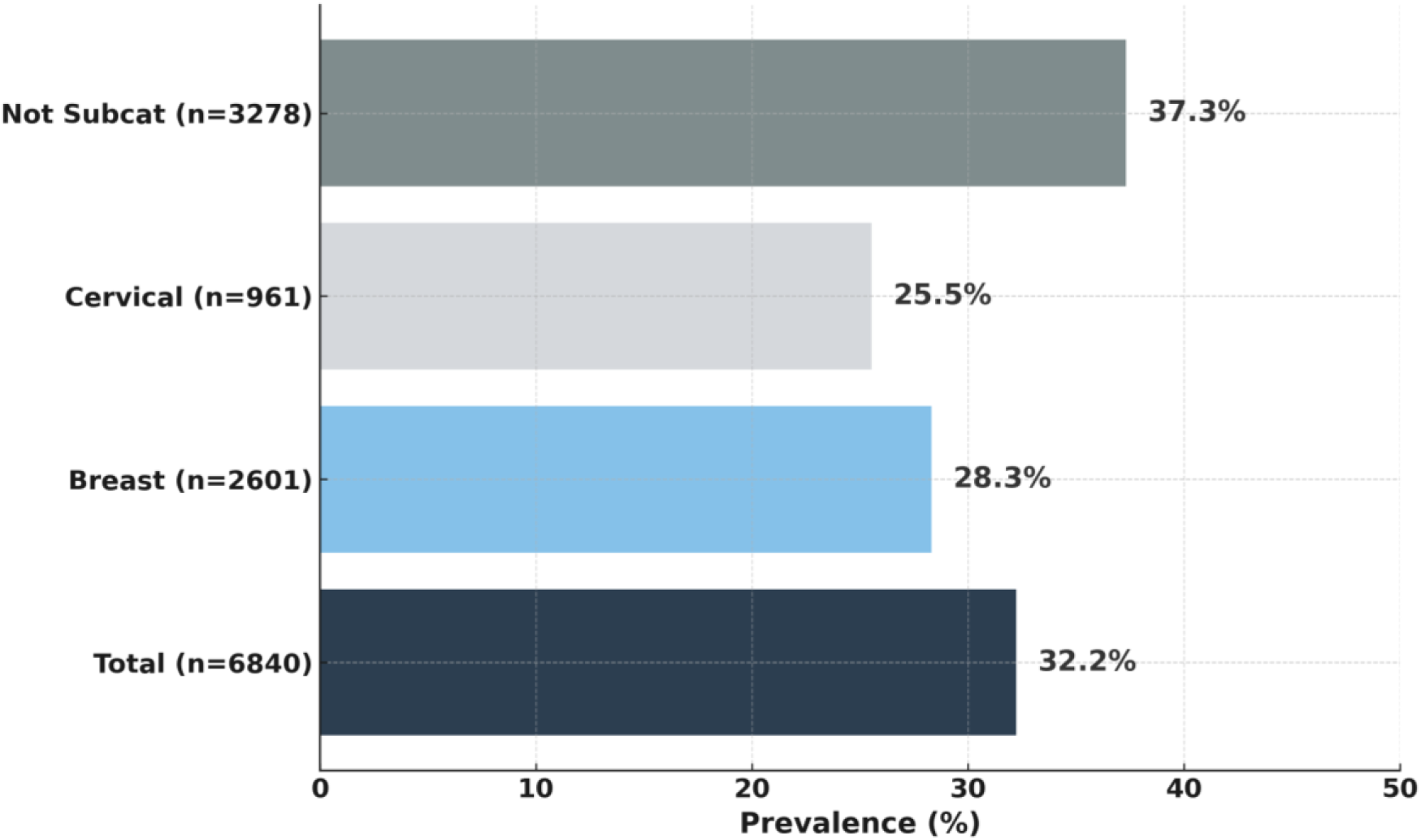
Pooled prevalence of IPV among women with cancer across six retrospective studies. The overall pooled prevalence was 32.2% (n=6,840). Subgroup analyses by cancer type show a prevalence of 28.3% for breast cancer (n=2,601), 25.5% for cervical cancer (n=961), and 37.3% in studies where the cancer type was not specified (n=3,278). Horizontal bars represent prevalence estimates, with labels indicating sample size.

**Table 5.** Characteristics of retrospective studies included in the pooled prevalence analysis IPV among women with cancer. The table details study authorship, year, journal, design, total sample size, cancer types considered, and IPV prevalence. When reported, both total and cancer-specific IPV prevalence rates are provided. This table forms the basis for the pooled estimates presented in Figure 3.

| Author | Year | Journal | Study Design | Sample Size (n) | Cancer Type(s) and Size | Reported IPV Prevalence |
| --- | --- | --- | --- | --- | --- | --- |
| Coker | 2017 A | Cancer Causes Control | Retrospective | 3278 | Not subcategorized | 37.3% |
| Jetelina | 2020 | J of Cancer Suvivorship | Retrospective, Population-based | 312 | Breast (257); Cervical (54) | TOT 54%; Breast 40.71%; Cervical 11.22% |
| Thananowan | 2016 | J of Interpersonal Violence | Cross-sectional | 532 | Cervical | 21.1% |
| Fouladi | 2021 | J of Caring Sciences | Retrospective | 211 | Breast | 90% |
| Baldwin | 2016 | Gynecologic Oncology | Population based (Kentuky and North Carolina Cancer Registry) | 2078 | Breast (1981); Cervical (97) | TOT 27.24%; Cervical 38.1%; Breast 26.7% |
| Meng | 2023 | Nursing Open | Cross-sectional | 429 | Cervical (218); Endometrial (70); Ovarian (104); Other (37) | TOT 31%; Cervical 28%; Endometrial 32.9%; Ovarian 34.6%; Other 35.1% |

## DISCUSSION AND LIMITATIONS OF THE STUDY

### State of the art and major findings

This systematic review was conducted to assess whether intimate partner violence (IPV) constitutes an independent risk factor for cancer, beyond its well-documented impact on women’s physical and mental health. To this end, we systematically synthesized available longitudinal and retrospective evidence specifically addressing the association between IPV and cancer diagnoses or clinically relevant precursor lesions, integrating quantitative analyses with a critical evaluation of potential biological and behavioral mechanisms.

To contextualize the present findings, it is important to consider prior attempts to address this research question. To date, the only published meta-analysis on the topic is the study by González et al.^17^ (2018). While pioneering, that work is limited by substantial methodological heterogeneity. Specifically, it combines disparate exposures—including adverse childhood experiences (ACEs), childhood abuse, and non-partner violence—with highly heterogeneous outcomes, ranging from cancer screening and adherence to diagnosis and mortality. Furthermore, the included studies vary considerably in quality, and adjusted estimates are inconsistently reported. As such, while the meta-analytic estimates reported by González et al. (e.g., OR = 2.54 for cervical cancer) are statistically significant, they lack clinical interpretability and do not permit inference regarding IPV as a distinct and independent risk factor for cancer development^17^.

In contrast, the present review employed stringent inclusion criteria, focusing exclusively on IPV perpetrated against women and limiting outcomes to confirmed cancer diagnoses or well-defined precursor conditions. Furthermore, only studies reporting formal measures of association adjusted for key confounding variables were eligible for inclusion in the meta-analysis. As a result, only two studies satisfied these criteria^15,16^, underscoring a fundamental limitation in the existing literature and precluding any definitive conclusions regarding a causal linkage.

Indeed, the primary conclusion emerging from our analysis is not the confirmation of an association, but rather the paucity of high-quality evidence addressing this question. Among the thirteen studies included in the review, only a minority were longitudinal^15,16,19,38^, and the majority employed cross-sectional or retrospective designs. Most investigations were restricted to cervical and breast cancer, with no studies examining other major malignancies such as colorectal, lung, or hematologic cancers. This narrow focus persists despite the biological plausibility of a more generalizable association, given the involvement of cancer-type-independent mechanisms such as chronic inflammation^44,45^, immune dysfunction^46–48^, and dysregulation of the hypothalamic– pituitary–adrenal axis^10,11,44,45,49,50^.

Although our pooled prevalence analyses and meta-analytic results may suggest a potential link between IPV and increased cancer burden, these findings must be interpreted with caution. The most salient result of this review is the structural absence of rigorous, prospective research on the oncologic consequences of IPV. The limited number of available studies, their methodological heterogeneity, and the absence of adequate control for confounding factors collectively prevent meaningful risk estimation or mechanistic inference. Addressing this gap should be considered a priority in both oncologic and public health research agendas.

The results of our meta-analysis and pooled prevalence analysis suggest a potential association between IPV and cancer, particularly cervical cancer. Among the few longitudinal studies available, Coker et al. (2000)¹⁵ reported a significantly increased risk of invasive cervical cancer in women with a history of IPV (aRR = 4.28), while Cesario et al.¹⁹ described a ten-fold higher prevalence of cervical cancer among IPV survivors. However, the overall number of longitudinal studies with formal risk estimates remains limited, and methodological differences preclude definitive conclusions. Retrospective studies provide additional insights, particularly regarding breast cancer. Jetelina et al.²⁰ identified a higher prevalence of aggressive ER-/PR- tumors in women with a history of IPV, along with disparities in access to surgery and hormonal therapy. These patterns highlight the possibility that IPV may affect not only cancer risk but also disease severity and treatment trajectories.

Our pooled analysis of retrospective studies (Figure 3, Table 5) showed that approximately 32.2% of women diagnosed with cancer report a lifetime history of IPV, with substantial rates in both breast (28.3%) and cervical cancer (25.5%) subgroups. These findings point to a potentially high burden of IPV among oncology patients and reinforce the need for trauma-informed models of care. Nonetheless, the available evidence does not yet permit a firm conclusion regarding a bidirectional or causal linkage between IPV and cancer development.

Cervical cancer emerges as the malignancy most consistently associated with IPV in the available literature^20,21,38^. This may reflect both biological vulnerability—due to increased risk of HPV exposure from coerced sexual activity, early sexual debut, and unprotected sex^38,51^—and structural barriers to preventive care. Several studies have reported lower participation in cervical cancer screening among IPV survivors, attributable to trauma, fear, and controlling partner behaviors^18,19,42^. These factors contribute to diagnostic delays and poorer prognosis, underscoring the urgent need for integrated IPV screening and trauma-sensitive gynecological care pathways^52,53^.

Emerging data on breast cancer also point toward a potential link with IPV, particularly in relation to more aggressive tumor phenotypes and reduced access to appropriate treatments^20,39^. Psychosocial barriers such as fear, mistrust, and economic instability may limit adherence to care, further exacerbating outcomes^54^. Taken together, these findings suggest that IPV may be an overlooked determinant of cancer risk and survivorship, with implications for both clinical management and public health policy^12^. Targeted prevention and screening strategies tailored to the needs of IPV survivors should be considered a critical priority^55^.

### Potential Mechanisms Linking IPV and Oncogenesis

Although the current evidence base does not allow for causal linkage, several converging lines of evidence point toward a plausible link between IPV and cancer risk. As discussed in the previous sections regarding the role of socioeconomic status, here we focus on biological and behavioral mechanisms that may help explain why an association between IPV and oncogenesis is plausible. In particular, the repeated association with cervical cancer, the high prevalence of IPV among women with cancer, and the presence of biologically plausible pathways suggest that the relationship may be underdetected rather than absent. Mechanistic hypotheses are therefore essential to guide future research, especially in identifying potential biological and behavioral mediators linking IPV to oncogenesis.

Among the most debilitating sequelae of intimate partner violence (IPV) are psychological disorders such as depression, anxiety, and post-traumatic stress disorder (PTSD). These conditions have well-established impacts on mental health and are increasingly recognized as contributors to adverse physical health outcomes, including cancer.^8,10,12,25,56^

Psychosocial stress, especially when chronic, has been associated with dysregulation of key biological systems involved in oncogenesis.^57,58^ Accumulating evidence links both acute and chronic stress to increased cancer risk and impaired immune surveillance.^59–62^ However, findings across studies remain inconsistent, likely due to methodological heterogeneity in exposure and outcome measures, study design, and control of confounders.^63–68^

Despite these inconsistencies, there is growing interest in PTSD as a potential etiological factor in cancer. PTSD disproportionately affects women, with prevalence rates nearly twice as high as those observed in men.^69^ Though underexplored, emerging evidence suggests a possible link between PTSD and increased cancer incidence and mortality. Roberts et al.^70^ reported elevated cancer-related mortality among women with PTSD, particularly from ovarian cancer—a finding supported by a recent meta-analysis.^71^ Behavioral pathways commonly associated with PTSD, including reduced adherence to gynecological screening,^72^ higher alcohol consumption,^73^ and increased rates of obesity,^74^ may mediate this association.

Depression, frequently comorbid with anxiety, also appears to influence cancer vulnerability and progression. It has been associated with increased risk of ovarian cancer^75^ and poorer outcomes in breast cancer, including reduced survival, higher recurrence, and increased all-cause mortality.^76,77^ Beyond individual psychopathology, psychosocial factors such as reduced emotional and social support have been linked to diminished survival in ovarian,^78^ breast,^79^ and colorectal cancer.^80^ Importantly, psychological interventions—such as supportive-expressive group therapy—may improve both mental health and survival in women with cancer, suggesting that psychosocial care can modulate clinical trajectories.^81^

These psychological and behavioral consequences of IPV are closely intertwined with biological mechanisms that may directly promote tumorigenesis. IPV has been associated with immune dysregulation, chronic inflammation, and endocrine disruption.^10,11,25,44,45,49,50^ For instance, C-reactive protein (CRP), a biomarker of systemic low-grade inflammation, is elevated not only in response to infection but also in states of chronic psychological stress such as PTSD and IPV.^46,82–85^ Persistent trauma-related arousal may lead to sustained immune activation, resulting in chronic inflammation.^47,48,84^ Elevated CRP levels have been associated with increased risk of various cancers—including gastrointestinal, hepatic, pulmonary, renal, and hematological malignancies^86^—and with cancer mortality in individuals with comorbid cardiovascular disease.^87^

IPV-related stress also impacts endocrine function. Dysregulation of the hypothalamic– pituitary–adrenal (HPA) axis in IPV survivors results in abnormal cortisol patterns, such as blunted diurnal variation or elevated evening levels.^25^ These disruptions are often accompanied by altered estradiol dynamics, impairing the regulation of emotional memory and fear extinction.^88,89^ Neuroendocrine changes extend to altered brain-derived neurotrophic factor (BDNF) signaling, aberrant circadian rhythms, and structural changes in brain regions including the hippocampus, amygdala, and prefrontal cortex—regions implicated in both stress adaptation and oncogenesis.^90–92^

Estrogen receptor signaling is a key pathway through which these hormonal disruptions may contribute to cancer development. Estrogen receptors (ERα and ERβ) mediate the effects of estradiol on cell proliferation, differentiation, and survival^93–95^ (Fig.4).

**Figure 4.**
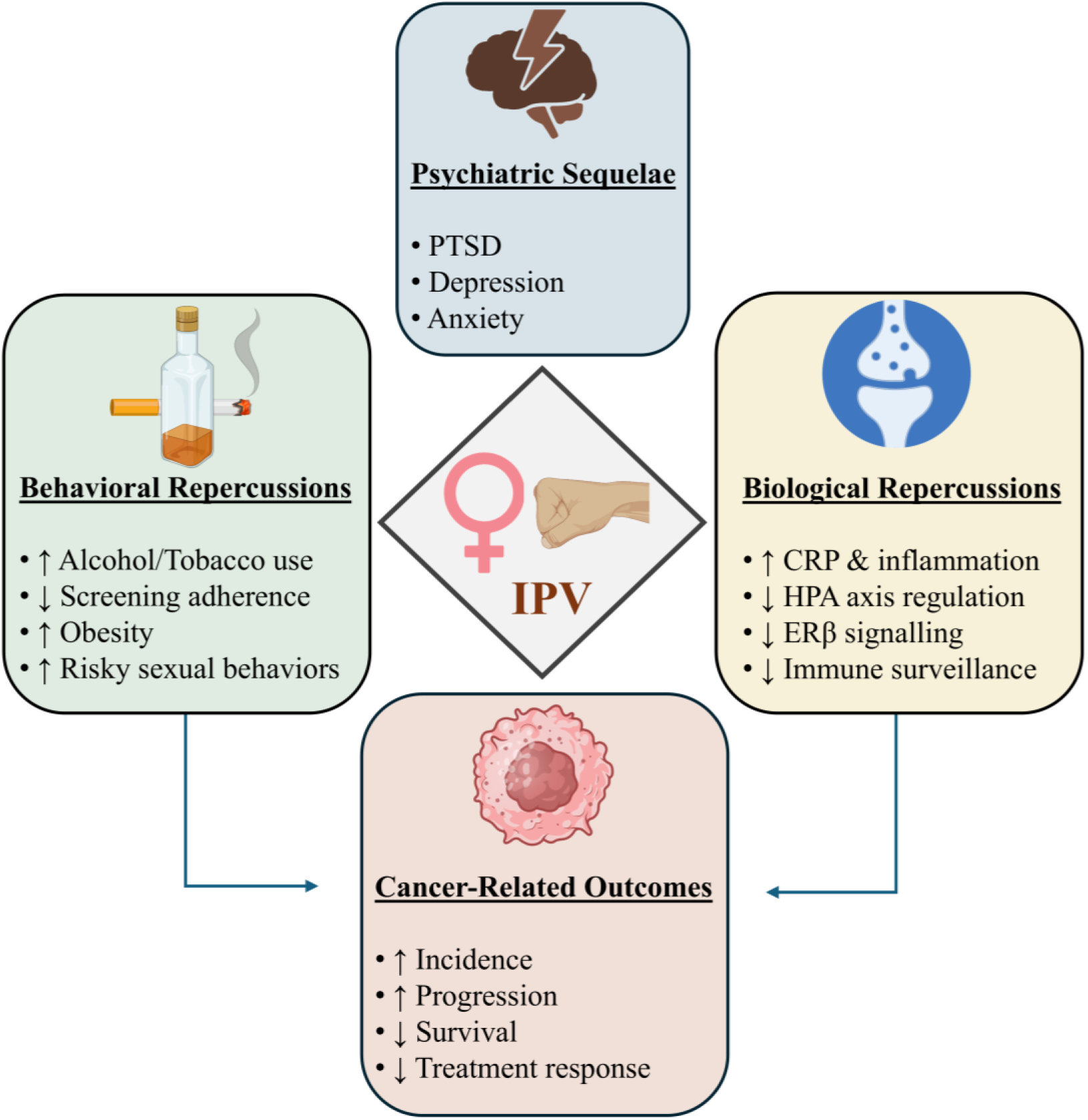
Conceptual framework illustrating potential mechanisms linking IPV exposure to cancer. The central diamond represents IPV exposure, which can initiate a cascade of psychiatric, behavioral, and biological repercussions. Psychiatric sequelae (e.g., PTSD, depression, anxiety, emotional dysregulation) may exacerbate behavioral alterations such as increased use of alcohol or tobacco, reduced adherence to cancer screening, obesity, and engagement in risky sexual behaviors. IPV-related biological disruptions include systemic inflammation (e.g., elevated CRP), hypothalamic-pituitary-adrenal (HPA) axis dysregulation, impaired estrogen receptor beta (ERβ) signaling, and reduced immune surveillance. Both behavioral and biological pathways may independently or synergistically increase cancer risk, influence tumor progression, reduce survival, and affect treatment response.

Their role extends beyond reproductive tissues to systems involved in metabolism, neurobiology, and immune regulation.^96,97^ In hormone-sensitive cancers—particularly breast, endometrial, and ovarian—alterations in estrogen signaling can critically affect tumor behavior and therapeutic response.^98–104^ ER-positive tumors typically respond to hormone therapy, improving survival,^101–103^ while ER-negative tumors are more aggressive and harder to treat.^104^ Emerging evidence from preclinical studies suggests that chronic stress and aggression associated with IPV may suppress ERβ signaling in the brain, particularly in the hippocampus, where this receptor plays a protective role in emotional regulation and neuronal plasticity.^105–107^ In rodent models of male-to-female violence, researchers have observed reduced ERβ expression, increased glucocorticoid levels, and downregulation of BDNF/TrkB signaling in the hippocampus.^108^ These findings point to a potential molecular mechanism through which chronic stress may alter neuroendocrine function and contribute to cancer vulnerability, although translational studies in humans are still lacking.

In sum, IPV may influence cancer risk and progression through a confluence of behavioral, psychosocial, and biological mechanisms (Fig.4). These include reduced access to preventive care, maladaptive coping behaviors, chronic inflammation, immune suppression, neuroendocrine disruption, and altered hormonal signaling. While the causal linkage remains incompletely understood, clarifying these mechanisms is essential to advancing both cancer prevention and trauma-informed care strategies for IPV survivors.

### Obstacles to Evidence-Based Integration of IPV in Cancer Research and Care

Despite the emerging evidence of an association between IPV and cancer, substantial gaps and limitations persist within the current body of research, limiting the capacity to fully understand and address this critical intersection.

First, a major limitation is the narrow focus on cervical cancer, with other cancer types receiving little to no attention. While cervical cancer is undoubtedly a critical endpoint, especially due to its link with HPV, other high-burden malignancies, such as breast, colorectal, lung, and hematologic cancers, have been largely overlooked. This is particularly problematic given that the biological mechanisms activated by IPV — including chronic stress, immune dysfunction, and behavioral risk factors — are relevant across multiple cancer types, not solely cervical cancer.^13,54,109^ Addressing this limitation requires studies that broaden the scope to include a wider variety of cancers, in order to capture the full extent of IPV’s impact on women’s health.

Second, there is a severe shortage of longitudinal studies that could establish causal linkage and temporal sequences between IPV and cancer development. Most available studies are cross-sectional or retrospective, thus preventing us from determining whether IPV precedes cancer onset, or whether women with cancer are at increased risk of experiencing IPV post-diagnosis. As noted by Thananowan et al.,^42^ longitudinal research is crucial to disentangle these relationships and to identify possible critical windows of vulnerability. Without prospective data, the role of IPV in cancer risk remains hypothetical, even though supported by biological plausibility and indirect evidence.

Third, many studies fail to adequately control for key confounders, including HPV infection, smoking, sexual behavior, substance use, socioeconomic status, and access to healthcare.^16,110^ HPV infection, in particular, is a major mediator of cervical cancer, and none of the studies included in our meta-analysis adjusted for HPV, thus complicating the interpretation of IPV as an independent risk factor. Similarly, behaviors like smoking and alcohol use, more frequent among IPV survivors,^4,54^ may confound the relationship with cancer if not properly accounted for. The lack of rigorous confounder adjustment weakens the strength of the evidence and highlights an essential methodological flaw.

Fourth, psychological, emotional, and economic forms of IPV are substantially underrepresented in current research, despite evidence that these forms of violence can have severe physiological consequences.^8^ The focus on physical and sexual violence neglects a critical dimension of IPV that could contribute to cancer risk through chronic stress, depression, and immune dysfunction.^42,108^ This represents a profound limitation, as psychological abuse often occurs in isolation or alongside other forms of IPV, and its biological impact is no less significant. Moreover, the heterogeneity in how IPV was defined and measured across the included studies further limits the comparability of findings. While we adopted the WHO definition of IPV as “any behavior within an intimate relationship that causes physical, psychological, or sexual harm,” many primary studies focused only on specific forms of IPV (e.g., physical or sexual violence), leaving psychological and emotional forms underrepresented. This variability highlights the need for future research to stratify analyses by type of IPV exposure (physical, sexual, psychological) to clarify whether different forms of violence exert distinct effects on cancer risk.

Fifth, geographic and cultural biases limit the generalizability of existing findings. The vast majority of studies have been conducted in Western, high-income countries,^54^ leaving a major gap in understanding IPV-related cancer risk in low- and middle-income countries (LMICs), where both IPV prevalence and barriers to healthcare access are often higher. This skews the evidence base and prevents a global understanding of IPV as a cancer risk factor. Finally, inconsistency in IPV definitions and measurement tools hinders comparability between studies. Many studies rely on abbreviated IPV measures or self-reports, which may underestimate prevalence due to stigma, fear, and recall bias.^8,22^ Moreover, diverse operationalizations of IPV — ranging from single-question assessments to complex validated tools — make it challenging to compare results and to develop standardized recommendations for screening and intervention. This variability also affects the validity of the observed associations, as different definitions may capture different dimensions and severities of IPV, with varying biological implications.

## CONCLUSIONS

This systematic review and meta-analysis reveal a striking paucity of high-quality studies examining the relationship between intimate partner violence (IPV) and cancer. Despite growing biological plausibility, only two longitudinal studies met inclusion criteria for meta-analysis, both focused exclusively on cervical cancer and based on overlapping populations. While a pooled odds ratio of 3.00 (95% CI: 2.05–4.38) was observed, the limited number of eligible studies prevents firm conclusions regarding causal linkage.

Our findings suggest a possible association between IPV and increased cancer risk, particularly for cervical and breast cancer, supported by preliminary evidence from retrospective studies and biologically plausible mechanisms involving chronic stress, immune dysfunction, and behavioral mediators. However, current evidence remains insufficient to fully characterize this relationship or to guide clinical decision-making.

There is a critical need for longitudinal, mechanistic, and population-based research to clarify the impact of IPV on cancer incidence and outcomes. Future studies should explore a wider range of cancer types, incorporate biomarkers of chronic stress and inflammation, and ensure adequate adjustment for confounders such as HPV infection and health-related behaviors. Moreover, validated tools for assessing IPV exposure—across its physical, psychological, and economic dimensions—are essential to improve research quality and comparability.

Recognizing IPV as a potential contributor to cancer risk could inform the development of targeted screening and prevention strategies for survivors. More broadly, integrating trauma-informed approaches into oncology and public health frameworks represents a necessary step toward addressing the long-term health consequences of violence against women.

## Data Availability

All data produced in the present study are available upon reasonable request to the authors

## RESOURSE AVAILABILITY

### Lead contact

Requests for further information and resources should be directed to and will be fulfilled by the lead contact, Jacopo Agrimi

### Materials availability

This study did not generate new unique reagents

### Data and code availability

This paper does not report original code. Data reported in this paper will be shared by the lead contact upon request.

## AUTHOR CONTRIBUTIONS

D.G.: investigation, writing, methodology

M.A.M.: investigation, writing, methodology

G.M.: investigation, writing

L.B.: investigation

N.P.: metodology, supervision, validation

M.S.: metodology, supervision, validation

J.A.: Conceptualisation, data curation, formal analysis, investigation, metodology, project administration, resources, software, supervision, validation, writing

G.S.: Conceptualisation, formal analysis, investigation, metodology, supervision, validation

## ACKNOWLEDGMENTS

Funding sources: JA, ACOM_UNCOVER_PRIN24-01

## REFERENCES

1. Organization WH. Intimate Partner Violence. 2022. https://apps.who.int/violence-info/intimate-partner-violence/ (accessed 03/01/2025 2025).

2. Krug EG, Mercy JA, Dahlberg LL, Zwi AB. [World report on violence and health]. Biomedica 2002; 22 **Suppl 2**: 327–36.

3. Jacquelyn C. Campbell KAK-T. Intimate Partner Violence: Implications for Women’s Physical and Mental Health. In: Kendall-Tackett KA, ed. Handbook of women stress and trauma: Brunner-Routledge; 2005.

4. Clemente-Teixeira M, Magalhaes T, Barrocas J, Dinis-Oliveira RJ, Taveira-Gomes T. Health Outcomes in Women Victims of Intimate Partner Violence: A 20-Year Real-World Study. Int J Environ Res Public Health 2022; 19(24).

5. Abramsky T, Watts CH, Garcia-Moreno C, et al. What factors are associated with recent intimate partner violence? findings from the WHO multi-country study on women’s health and domestic violence. BMC Public Health 2011; 11: 109.

6. Patra P, Prakash J, Patra B, Khanna P. Intimate partner violence: Wounds are deeper. Indian J Psychiatry 2018; 60(4): 494–8.

7. Mazza M, Marano G, Del Castillo AG, et al. Intimate partner violence: A loop of abuse, depression and victimization. World J Psychiatry 2021; 11(6): 215–21.

8. Stubbs A, Szoeke C. The Effect of Intimate Partner Violence on the Physical Health and Health-Related Behaviors of Women: A Systematic Review of the Literature. Trauma Violence Abuse 2022; 23(4): 1157–72.

9. Stockman JK, Hayashi H, Campbell JC. Intimate Partner Violence and its Health Impact on Ethnic Minority Women [corrected]. J Womens Health (Larchmt) 2015; 24(1): 62–79.

10. Campbell JC. Health consequences of intimate partner violence. Lancet 2002; 359(9314): 1331-6.

11. Campbell J, Jones AS, Dienemann J, et al. Intimate partner violence and physical health consequences. Arch Intern Med 2002; 162(10): 1157–63.

12. Bacchus LJ, Ranganathan M, Watts C, Devries K. Recent intimate partner violence against women and health: a systematic review and meta-analysis of cohort studies. BMJ Open 2018; 8(7): e019995.

13. Goldberg X. Female Survivors of Intimate Partner Violence (IPV) and Mental Health.; 2023.

14. Exner-Cortens D, Eckenrode J, Rothman E. Longitudinal associations between teen dating violence victimization and adverse health outcomes. Pediatrics 2013; 131(1): 71–8.

15. Coker AL, Sanderson M, Fadden MK, Pirisi L. Intimate partner violence and cervical neoplasia. J Womens Health Gend Based Med 2000; 9(9): 1015–23.

16. Coker AL, Hopenhayn C, DeSimone CP, Bush HM, Crofford L. Violence against Women Raises Risk of Cervical Cancer. J Womens Health (Larchmt) 2009; 18(8): 1179–85.

17. Reingle Gonzalez JM, Jetelina KK, Olague S, Wondrack JG. Violence against women increases cancer diagnoses: Results from a meta-analytic review. Prev Med 2018; 114: 168–79.

18. Bagwell-Gray ME, Ramaswamy M. Cervical Cancer Screening and Prevention among Survivors of Intimate Partner Violence. Health Soc Work 2022; 47(2): 102–12.

19. Cesario SK, McFarlane J, Nava A, Gilroy H, Maddoux J. Linking cancer and intimate partner violence: the importance of screening women in the oncology setting. Clin J Oncol Nurs 2014; 18(1): 65–73.

20. Jetelina KK, Carr C, Murphy CC, Sadeghi N, J SL, Tiro JA. The impact of intimate partner violence on breast and cervical cancer survivors in an integrated, safety-net setting. J Cancer Surviv 2020; 14(6): 906–14.

21. Hindin PB, R.; Brown, D.R.; Munet-Vilaro, F. Intimate Partner Violence and Risk for Cervical Cancer. Journal of Family Violence 2015; 30: 1031–43.

22. Coker AL. Does Physical Intimate Partner Violence Affect Sexual Health? Trauma, Violence, & Abuse 2007; 8(2): 149–77.

23. Liu Y, Tian S, Ning B, Huang T, Li Y, Wei Y. Stress and cancer: The mechanisms of immune dysregulation and management. Front Immunol 2022; 13: 1032294.

24. Lei Y, Liao F, Tian Y, Wang Y, Xia F, Wang J. Investigating the crosstalk between chronic stress and immune cells: implications for enhanced cancer therapy. Front Neurosci 2023; 17: 1321176.

25. Cesari V, Vallefuoco A, Agrimi J, Gemignani A, Paolocci N, Menicucci D. Intimate partner violence: psycho-physio-pathological sequelae for defining a holistic enriched treatment. Front Behav Neurosci 2022; 16: 943081.

26. Agrimi J, Bernardele L, Sbaiti N, et al. Male violence disrupts estrogen receptor beta signaling in the female hippocampus. bioRxiv 2023.

27. Page MJ, McKenzie JE, Bossuyt PM, et al. The PRISMA 2020 statement: an updated guideline for reporting systematic reviews. BMJ 2021: n71.

28. Brooke BS, Schwartz TA, Pawlik TM. MOOSE Reporting Guidelines for Meta-analyses of Observational Studies. JAMA Surgery 2021; 156(8): 787.

29. Solomon D. The 2001 Bethesda System Terminology for Reporting Results of Cervical Cytology. JAMA 2002; 287(16): 2114.

30. Beel J, Gipp B. Google Scholar’s Ranking Algorithm: An Introductory Overview. Proceedings of ISSI 2009 2009: 230–41.

31. Haddaway NR, Collins AM, Coughlin D, Kirk S. The Role of Google Scholar in Evidence Reviews and Its Applicability to Grey Literature Searching. PLoS One 2015; 10(9): e0138237.

32. Bramer WM, Rethlefsen ML, Kleijnen J, Franco OH. Optimal database combinations for literature searches in systematic reviews: a prospective exploratory study. Syst Rev 2017; 6(1): 245.

33. Coker AL, Follingstad DR, Garcia LS, Bush HM. Intimate partner violence and women’s cancer quality of life. Cancer Causes Control 2017; 28(1): 23–39.

34. Coker AL, Follingstad DR, Garcia LS, Bush HM. Partner interfering behaviors affecting cancer quality of life. Psychooncology 2017; 26(8): 1205–14.

35. Coker AL, Follingstad D, Garcia LS, Williams CM, Crawford TN, Bush HM. Association of intimate partner violence and childhood sexual abuse with cancer-related well-being in women. J Womens Health (Larchmt) 2012; 21(11): 1180–8.

36. Devries KM, Mak JY, Bacchus LJ, et al. Intimate partner violence and incident depressive symptoms and suicide attempts: a systematic review of longitudinal studies. PLoS Med 2013; 10(5): e1001439.

37. Yount KM, Crandall A, Cheong YF, et al. Child Marriage and Intimate Partner Violence in Rural Bangladesh: A Longitudinal Multilevel Analysis. Demography 2016; 53(6): 1821–52.

38. Madding RA, Currier JJ, Yanit K, Hedges M, Bruegl A. HPV self-collection for cervical cancer screening among survivors of sexual trauma: a qualitative study. BMC Womens Health 2024; 24(1): 509.

39. Fouladi N, Feizi I, Pourfarzi F, et al. Factors Affecting Behaviors of Women with Breast Cancer Facing Intimate Partner Violence Based on PRECEDE-PROCEED Model. J Caring Sci 2021; 10(2): 89–95.

40. Baldwin LA, Roberts ME, Lefringhouse J, et al. Frequency of intimate partner violence history in gynecologic and breast cancers. Gynecologic Oncology 2016; 141: 124.

41. Ramaswamy M, Kelly PJ, Koblitz A, Kimminau KS, Engelman KK. Understanding the role of violence in incarcerated women’s cervical cancer screening and history. Women Health 2011; 51(5): 423–41.

42. Thananowan N, Vongsirimas N. Factors Mediating the Relationship Between Intimate Partner Violence and Cervical Cancer Among Thai Women. J Interpers Violence 2016; 31(4): 715–31.

43. Meng Y, Shang M, Cai T, et al. Incidence and risk factors of intimate partner violence among patients with gynaecological cancer in China. Nurs Open 2023; 10(8): 5338–47.

44. Black PH. The inflammatory consequences of psychologic stress: Relationship to insulin resistance, obesity, atherosclerosis and diabetes mellitus, type II. Medical Hypotheses 2006; 67(4): 879–91.

45. McEwen BS. Neurobiological and Systemic Effects of Chronic Stress. Chronic Stress 2017; 1: 247054701769232.

46. Spitzer C, Wibisono D, Terfehr K, Löwe B, Otte C, Wingenfeld K. C-reactive protein, pre- and postdexamethasone cortisol levels in post-traumatic stress disorder. Nordic Journal of Psychiatry 2014; 68(5): 296–9.

47. Du Clos TW, Mold C. C-Reactive Protein: An Activator of Innate Immunity and a Modulator of Adaptive Immunity. Immunologic Research 2004; 30(3): 261–78.

48. Hassamal S. Chronic stress, neuroinflammation, and depression: an overview of pathophysiological mechanisms and emerging anti-inflammatories. Frontiers in Psychiatry 2023; 14.

49. Davis T. Cortisol Dysregulation and the Trauma Cycle in Intimate Partner Violence Survivors. Journal of Clinical Psychology and Neurology 2025; 3: 1–8.

50. Potter LC, Morris R, Hegarty K, García-Moreno C, Feder G. Categories and health impacts of intimate partner violence in the World Health Organization multi-country study on women’s health and domestic violence. International Journal of Epidemiology 2021; 50(2): 652–62.

51. Calvillo C, Marshall A, Gafford S, Montgomery BEE. Intimate partner violence and its relation to sexual health outcomes across different adult populations: a systematic review. Frontiers in Sociology 2024; 9.

52. Amanda LT, Stephanie B, Rishi R, et al. Screening and intervention for intimate partner violence at trauma centers and emergency departments: an evidence-based systematic review from the Eastern Association for the Surgery of Trauma. Trauma Surgery & Acute Care Open 2023; 8(1): e001041.

53. Crighton D, Towl G. Meta-analysis suggests trauma informed care for women subjected to intimate partner violence (IPV) may be effective in reducing depression and anxiety in survivors. Evidence Based Nursing 2025; 28(1): 15.

54. Goldberg X, Espelt C, Porta-Casteràs D, Palao D, Nadal R, Armario A. Non-communicable diseases among women survivors of intimate partner violence: Critical review from a chronic stress framework. Neuroscience & Biobehavioral Reviews 2021; 128: 720–34.

55. Phares TM, Sherin K, Harrison SL, Mitchell C, Freeman R, Lichtenberg K. Intimate Partner Violence Screening and Intervention: The American College of Preventive Medicine Position Statement. Am J Prev Med 2019; 57(6): 862–72.

56. 56. Cristea IA, Stefan S, David O, Mogoase C, Dobrean A. Rational-Emotive and Cognitive-Behavior Therapy for Generalized Anxiety Disorder. Springer International Publishing; 2016: 15-30.

57. Moreno-Smith M, Lutgendorf SK, Sood AK. Impact of Stress on Cancer Metastasis. Future Oncology 2010; 6(12): 1863–81.

58. Dai S, Mo Y, Wang Y, et al. Chronic Stress Promotes Cancer Development. Front Oncol 2020; 10: 1492.

59. Lillberg K. Stressful Life Events and Risk of Breast Cancer in 10,808 Women: A Cohort Study. American Journal of Epidemiology 2003; 157(5): 415–23.

60. Helgesson O, Cabrera C, Lapidus L, Bengtsson C, Lissner L. Self-reported stress levels predict subsequent breast cancer in a cohort of Swedish women. Eur J Cancer Prev 2003; 12(5): 377–81.

61. Palesh O, Butler LD, Koopman C, Giese-Davis J, Carlson R, Spiegel D. Stress history and breast cancer recurrence. Journal of Psychosomatic Research 2007; 63(3): 233–9.

62. Chiriac VF, Baban A, Dumitrascu DL. Psychological stress and breast cancer incidence: a systematic review. Clujul Med 2018; 91(1): 18–26.

63. Nielsen NR, Zhang Z-F, Kristensen TS, Netterstr⊘M B, Schnohr P, Gr⊘Nbæk M. Self reported stress and risk of breast cancer: prospective cohort study. BMJ 2005; 331(7516): 548.

64. Heikkila K, Nyberg ST, Theorell T, et al. Work stress and risk of cancer: meta-analysis of 5700 incident cancer events in 116,000 European men and women. BMJ 2013; 346: f165.

65. Santos MCL, Horta BL, Amaral JJFD, Fernandes PFCBC, Galvão CM, Fernandes AFC. Association between stress and breast cancer in women: a meta-analysis. Cadernos de Saúde Pública 2009; 25(suppl 3): S453–S63.

66. Trudel-Fitzgerald C, Poole EM, Idahl A, et al. The Association of Work Characteristics With Ovarian Cancer Risk and Mortality. Psychosomatic Medicine 2017; 79(9): 1059–67.

67. Garssen B. Psychological factors and cancer development: evidence after 30 years of research. Clin Psychol Rev 2004; 24(3): 315–38.

68. Eckerling A, Ricon-Becker I, Sorski L, Sandbank E, Ben-Eliyahu S. Stress and cancer: mechanisms, significance and future directions. Nat Rev Cancer 2021; 21(12): 767–85.

69. Kessler RC. Posttraumatic Stress Disorder in the National Comorbidity Survey. Archives of General Psychiatry 1995; 52(12): 1048.

70. Roberts AL, Huang T, Koenen KC, Kim Y, Kubzansky LD, Tworoger SS. Posttraumatic Stress Disorder Is Associated with Increased Risk of Ovarian Cancer: A Prospective and Retrospective Longitudinal Cohort Study. Cancer Research 2019; 79(19): 5113–20.

71. Yang J, Jiang W. A meta-analysis of the association between post-traumatic stress disorder and cancer risk. Frontiers in Psychiatry 2023; 14.

72. Cadman L, Waller J, Ashdown-Barr L, Szarewski A. Barriers to cervical screening in women who have experienced sexual abuse: an exploratory study. J Fam Plann Reprod Health Care 2012; 38(4): 214–20.

73. Danielson CK, Amstadter AB, Dangelmaier RE, Resnick HS, Saunders BE, Kilpatrick DG. Trauma-related risk factors for substance abuse among male versus female young adults. Addict Behav 2009; 34(4): 395–9.

74. Perkonigg A, Owashi T, Stein MB, Kirschbaum C, Wittchen H-U. Posttraumatic Stress Disorder and Obesity. American Journal of Preventive Medicine 2009; 36(1): 1–8.

75. Huang T, Poole EM, Okereke OI, et al. Depression and risk of epithelial ovarian cancer: Results from two large prospective cohort studies. Gynecol Oncol 2015; 139(3): 481–6.

76. Wang Y-H, Li J-Q, Shi J-F, et al. Depression and anxiety in relation to cancer incidence and mortality: a systematic review and meta-analysis of cohort studies. Molecular Psychiatry 2020; 25(7): 1487–99.

77. Wang X, Wang N, Zhong L, et al. Prognostic value of depression and anxiety on breast cancer recurrence and mortality: a systematic review and meta-analysis of 282,203 patients. Molecular Psychiatry 2020; 25(12): 3186–97.

78. Lutgendorf SK, De Geest K, Bender D, et al. Social Influences on Clinical Outcomes of Patients With Ovarian Cancer. Journal of Clinical Oncology 2012; 30(23): 2885–90.

79. Chou AF, Stewart SL, Wild RC, Bloom JR. Social support and survival in young women with breast carcinoma. Psychooncology 2012; 21(2): 125–33.

80. Kroenke CH, Paskett ED, Cené CW, et al. Prediagnosis social support, social integration, living status, and colorectal cancer mortality in postmenopausal women from the women’s health initiative. Cancer 2020; 126(8): 1766–75.

81. Giese-Davis J, Collie K, Rancourt KM, Neri E, Kraemer HC, Spiegel D. Decrease in depression symptoms is associated with longer survival in patients with metastatic breast cancer: a secondary analysis. J Clin Oncol 2011; 29(4): 413–20.

82. Spitzer C, Barnow S, Völzke H, et al. Association of posttraumatic stress disorder with low-grade elevation of C-reactive protein: Evidence from the general population. Journal of Psychiatric Research 2010; 44(1): 15–21.

83. Luan Y-Y, Yao Y-M. The Clinical Significance and Potential Role of C-Reactive Protein in Chronic Inflammatory and Neurodegenerative Diseases. Frontiers in Immunology 2018; 9.

84. Heath NM, Chesney SA, Gerhart JI, et al. Interpersonal violence, PTSD, and inflammation: Potential psychogenic pathways to higher C-reactive protein levels. Cytokine 2013; 63(2): 172–8.

85. Powers A, Dixon HD, Conneely K, et al. The differential effects of PTSD, MDD, and dissociation on CRP in trauma-exposed women. Comprehensive Psychiatry 2019; 93: 33–40.

86. Zhu M, Ma Z, Zhang X, et al. C-reactive protein and cancer risk: a pan-cancer study of prospective cohort and Mendelian randomization analysis. BMC Medicine 2022; 20(1).

87. Endo H, Dohi T, Funamizu T, et al. Long-Term Predictive Value of High-Sensitivity C-Reactive Protein for Cancer Mortality in Patients Undergoing Percutaneous Coronary Intervention. Circulation Journal 2019; 83(3): 630–6.

88. Fries GR, Vasconcelos-Moreno MP, Gubert C, et al. Hypothalamic-Pituitary-Adrenal Axis Dysfunction and Illness Progression in Bipolar Disorder. International Journal of Neuropsychopharmacology 2014; 18(1).

89. Kinlein SA, Wilson CD, Karatsoreos IN. Dysregulated Hypothalamic–Pituitary– Adrenal Axis Function Contributes to Altered Endocrine and Neurobehavioral Responses to Acute Stress. Frontiers in Psychiatry 2015; **Volume 6 -** 2015.

90. Tsimpolis A, Kalafatakis K, Charalampopoulos I. Recent advances in the crosstalk between the brain-derived neurotrophic factor and glucocorticoids. Frontiers in Endocrinology 2024; **Volume 15 -** 2024.

91. Dou S-H, Cui Y, Huang S-M, Zhang B. The Role of Brain-Derived Neurotrophic Factor Signaling in Central Nervous System Disease Pathogenesis. Frontiers in Human Neuroscience 2022; **Volume 16 -** 2022.

92. Sequeira MK, Bolton JL. Stressed Microglia: Neuroendocrine–Neuroimmune Interactions in the Stress Response. Endocrinology 2023; 164(7).

93. Hua H, Zhang H, Kong Q, Jiang Y. Mechanisms for estrogen receptor expression in human cancer. Experimental Hematology & Oncology 2018; 7(1).

94. Pakdel F. Molecular Pathways of Estrogen Receptor Action. International Journal of Molecular Sciences 2018; 19(9): 2591.

95. Ventura-Clapier R, Piquereau J, Veksler V, Garnier A. Estrogens, Estrogen Receptors Effects on Cardiac and Skeletal Muscle Mitochondria. Frontiers in Endocrinology 2019; 10.

96. Den Ruijter HM, Kararigas G. Estrogen and Cardiovascular Health. Frontiers in Cardiovascular Medicine 2022; 9.

97. Willemars MMA, Nabben M, Verdonschot JAJ, Hoes MF. Evaluation of the Interaction of Sex Hormones and Cardiovascular Function and Health. Current Heart Failure Reports 2022; 19(4): 200–12.

98. Miziak P, Baran M, Błaszczak E, et al. Estrogen Receptor Signaling in Breast Cancer. Cancers 2023; 15(19): 4689.

99. Huang D, Huang Z, Indukuri R, et al. Estrogen Receptor β (ESR2) Transcriptome and Chromatin Binding in a Mantle Cell Lymphoma Tumor Model Reveal the Tumor-Suppressing Mechanisms of Estrogens. Cancers 2022; 14(13): 3098.

100. Chen H-L, Huang F-B, Chen Q, Deng Y-C. Impact of estrogen receptor expression level on response to neoadjuvant chemotherapy and prognosis in HER2-negative breast cancers. BMC Cancer 2023; 23(1).

101. Putti TC, El-Rehim DMA, Rakha EA, et al. Estrogen receptor-negative breast carcinomas: a review of morphology and immunophenotypical analysis. Modern Pathology 2005; 18(1): 26–35.

102. Ran R, Zhao S, Zhou Y, et al. Clinicopathological characteristics, treatment patterns and outcomes in patients with HER2-positive breast cancer based on hormone receptor status: a retrospective study. BMC Cancer 2024; 24(1).

103. Moy B, Wolff AC, Rumble RB, Allison KH, Carey LA. Chemotherapy and Targeted Therapy for Endocrine-Pretreated or Hormone Receptor–Negative Metastatic Breast Cancer and Human Epidermal Growth Factor Receptor 2 Testing in Breast Cancer: ASCO Guideline Rapid Recommendation Update Q and A. JCO Oncology Practice 2023; 19(8): 547–50.

104. Zabrocka E, Newman B, Levey G, et al. Estrogen Receptor-negative Ductal Carcinoma *In Situ*(DCIS) of the Breast – an Institutional Review of Outcomes. Anticancer Research 2023; **43**(9): 4031-6.

105. Zhao W, Hou Y, Zhang Q, et al. Estrogen receptor β exerts neuroprotective effects by fine-tuning mitochondrial homeostasis through NRF1/PGC-1α. Neurochemistry International 2023; 171: 105636.

106. Liu F, Day M, Muñiz LC, et al. Activation of estrogen receptor-β regulates hippocampal synaptic plasticity and improves memory. Nature Neuroscience 2008; 11(3): 334–43.

107. Priyanka HP, Pratap UP, Nair RS, Vasantharekha R, Thyagarajan S. Estrogen-receptor status determines differential regulation of α1- and α2-adrenoceptor-mediated cell survival, angiogenesis, and intracellular signaling responses in breast cancer cell lines. Medical Oncology 2024; 41(5).

108. Agrimi J, Bernardele L, Sbaiti N, et al. Reiterated male-to-female violence disrupts hippocampal estrogen receptor β expression, prompting anxiety-like behavior. iScience 2024; 27(9): 110585.

109. Zhu J, Exner-Cortens D, Dobson K, Wells L, Noel M, Madigan S. Adverse childhood experiences and intimate partner violence: A meta-analysis. Development and Psychopathology 2024; 36(2): 929–43.

110. McClintock HF, Dulak SL. Intimate Partner Violence and Sexually Transmitted Infections Among Women in Sub-Saharan Africa. Journal of Immigrant and Minority Health 2021; 23(2): 191–8.

